# Occurrence, fate, and risk assessment of antibiotics in typical pharmaceutical manufactories and receiving surface waters from different regions

**DOI:** 10.1101/2022.06.22.22276743

**Authors:** Yuanfei Liu, Dan Cai, Xin Li, Qingyao Wu, Ping Ding, Liangchen Shen, Jian Yang, Guocheng Hu, Jinhua Wu

**Affiliations:** School of Environment and Energy, South China University of Technology, Guangzhou, 510006, China; State Environmental Protection Key Laboratory of Environmental Pollution Health Risk Assessment, South China Institute of Environmental Sciences, Ministry of Ecology and Environment, Guangzhou, 510655, China; School of Public Health and Emergency Management, South University of Science and Technology of China, Shenzhen, 518055, China

**Keywords:** Antibiotics, Pharmaceutical plants, Receiving waters, Removal efficiency, Risk assessment

## Abstract

This study aimed to update information on the presence and persistence of antibiotics in wastewater from four typical pharmaceutical plants in China and the removal of antibiotics by the wastewater treatment process. It also evaluated the environmental impact of antibiotic residues through wastewater discharge into receiving water bodies. The results indicated that 13 antibiotics were detected in wastewater samples with concentrations ranging from 57.03 to 726.79 ng/L. Fluoroquinolones (FQs) and macrolides (MLs) were the most abundant antibiotic classes in wastewater samples, accounting for 42.5% and 38.7% of total antibiotic concentrations, respectively, followed by sulfonamides (SAs) (16.4%) and tetracyclines (TCs) (2.4%). Erythromycin-H_2_O (ERY), lincomycin (LIN), ofloxacin (OFL), and trimethoprim (TMP) were the most frequently detected antibiotics; among these antibiotics, the concentration of OFL was the highest in most wastewater samples. No significant difference was found in the removal of antibiotics between different treatment processes, and more than 50% of antibiotics were not completely removed with a removal efficiency of less than 70%. The concentration of detected antibiotics in the receiving water was an order of magnitude lower than that in the wastewater due to dilution. Finally, an environmental risk analysis showed that lincomycin and ofloxacin could pose a high risk at the concentrations detected in effluents and a medium risk in their receiving waters, highlighting that they were a potential hazard to the health of the aquatic ecosystem.

## 1. Introduction

Antibiotics are natural, synthetic, or semi-synthetic compounds that can kill or inhibit the growth or metabolic activity of microorganisms. These compounds are biologically active molecules with antibacterial, antifungal, and antiparasitic activities. They have been widely used to treat infectious diseases for both humans and animals in livestock farming, aquaculture, and agriculture (Anh et al., 2021). The data from 76 countries showed that the total global antibiotic consumption increased from 21.1 to 34.8 billion defined daily doses between 2000 and 2015 (Klein et al., 2018). Studies have shown that antibiotics are detected in surface water (Hirsch et al., 1999; Yan et al., 2013), groundwater (Barnes et al., 2008; Batt et al., 2006), domestic sewage (Andreozzi et al., 2004; Fatta-Kassinos et al., 2011; Gulkowska et al., 2008), sediment (Zhou et al., 2011), soil (Hu et al., 2010; Karci and Balcioǧlu, 2009), and even drinking water (Benotti et al., 2009), indicating that the environmental antibiotic pollution is widespread. In addition, the concentrations of antibiotics in Asian developing countries tend to be higher than those generally reported in European and North American countries (Kovalakova et al., 2020). It is estimated that more than 70 antibiotics with concentrations up to several micrograms per liter have been detected in 7 major water systems in China (Yin et al., 2021). Antibiotics in the environment may induce the production of antibiotic-resistant genes and spread of antibiotic-resistant bacteria and enter the human body through direct contact or via the food chain, leading to the development of resistance to some antibiotics (Hu et al., 2010; Zhang et al., 2015). Therefore, a comprehensive understanding of the environmental emission and fate and risk of antibiotics in waters is essential.

The primary sources of antibiotics in the environment are municipal wastewater treatment plants, agricultural settings, aquaculture, hospitals, and pharmaceutical production facilities (Kovalakova et al., 2020). Pharmaceutical manufactories have been proposed as important reservoirs of antibiotics. In most cases, the wastewater is discharged after treatment in the wastewater treatment facility of PMFs. However, antibiotic residues cannot be removed completely by the existing wastewater treatment process of PMFs.Research in Taiwan found that PMFs were an important source of antibiotics, with sulfamethoxazole having a maximum concentration exceeding 1,000,000 ng/L (Lin and Tsai, 2009). Pakistan reported a high level of antibiotic residues in wastewaters close to the pharmaceutical factories and a positive correlation between the level of residues and antibiotic resistance in the samples (Khan et al., 2013). Many researches have studied that the concentration levels of antibiotics detected in STPs might threaten the nontarget organisms, such as algae, *Daphnia magna*, mollusks, and fish (Białk-Bielińska et al., 2011; Gust et al., 2013; Isidori et al., 2005; Tkaczyk et al., 2021). However, current studies have mostly focused on removing antibiotics from sewage treatment plants (STPs) or the determination of antibiotics in natural water bodies, the occurrence, fate and environmental risk of antibiotics during wastewater treatment processes at PMFs are not well understood yet (Wang et al., 2020).

China’s pharmaceutical industry is developing rapidly and has grown to be the second largest pharmaceutical market in the world (Zhong and Ouyang, 2020), reaching 373 billion dollar in 2018. And recent review has reported the occurrence of a variety of antibiotics in the water environment in China (Qiao et al., 2018). Therefore, a systemic investigation of antibiotics in wastewaters originated from pharmaceutical manufacturing and the occurrence, fate, environmental risk assessment of antibiotics between typical PMFs and their receiving water in China is necessary. In the light of these concerns, the aim of the present study was to aimed to determine and monitor the occurrence of antibiotics in municipal wastewaters from 4 typical PMFs of 4 provinces in China.The presence of antibiotics were also monitored in their receiving water bodies, from upstream of the river to the wastewater discharge point and downstream of the discharge point, to assess the occurrence and impact of these antibiotics in these rivers. Finally, the potential ecological risks of the target antibiotics to aquatic species were assessed according to the calculated risk quotients, and the results provided important background data for the pollution control of antibiotics in the aquatic environment in these areas.

## 2. Materials and Methods

### 2.1. Materials and reagents

The 17 target antibiotics belonging to 5 different classes included the following: (I) 1 TC, metacycline (MTC); (II) 9 SAs, sulfachlorpyridazine (SCP), sulfadiazine (SDZ), sulfadimethoxine (SDM), sulfamethazine (SMZ), sulfameter (SME), sulfamethoxazole (SMX), sulfamonomethoxine (SMM), sulfapyridine (SPD), and trimethoprim (TMP); (III) 4 MLs, clarithromycin (CTM), erythromycin-H_2_O (ERY), LIN, and roxithromycin (ROX); and (IV) 3 FQs, ciprofloxacin (CIP), norfloxacin (NFX), and ofloxacin (OFL), All the antibiotics standards were of high purity grade [high-performance liquid chromatography (HPLC) grade, >99%] and purchased from Anpel (Shanghai, China). Isotopically labeled compounds, including sulfamethoxazole-D_4_ (SMX-D_4_), erythromycin-^13^C-D_3_ (ERY-^13^C-D_3_), thiabendazole-D_4_ (TBD-D_4_), ciprofloxacin-D_8_ (CFX-D_8_), sulfamethazine-^13^C_6_ (SMZ-^13^C_6_), trimethoprim-D_3_ (TMP-D_3_) and lincomycin-D_3_ (LIN-D_3_), were obtained from Cambridge Isotope Laboratories (England) and C/D/N Isotopes (Canada).

Reagent-grade methanol, acetonitrile, formic acid, and other chemicals were purchased from local suppliers. Milli-Q water was used throughout the study. The stock solution of each antibiotic class was prepared in methanol. Working solutions with different concentrations were prepared by mixing and diluting the stock solutions.

### 2.2 Sample collection and preparation

The wastewater samples were collected from the influent and effluent of four PMFs, as well as their receiving water bodies in Hebei, Jiangsu, Zhejiang and Guangdong province of China. The specific sampling locations are depicted in Figure 1. Eight sampling points are georeferenced in Table S1. The basic information of each PMF is shown in Table S2. Different sewage treatment technologies used in the four PMFs included A2/O (Anaerobic-Anoxic-Oxic), MBR (Membrane Bio-Reactor), modified A/O (Anaerobic-Oxic), BAF (Biological Aerated Filters). The process flow charts and sampling sites are shown in Figure 2. Wastewater samples were collected from the influent and effluent of PMFs. Besides, the surface water from the upstream/downstream of the receiving rivers (100 m away from the effluent outfall) was also collected. All the samples in three replicates were obtained during November–December 2020 and July 2021. The samples from the PMFs were collected in 1-L amber glass as 24-h composite, while the samples from receiving rivers were collected as grab samples in the middle of the day. All samples were placed in a cool place at –20°C and analyzed within 7 days to minimize degradation. Each analysis was repeated three times, and the reported results were based on the average value.

**Figure 1.**
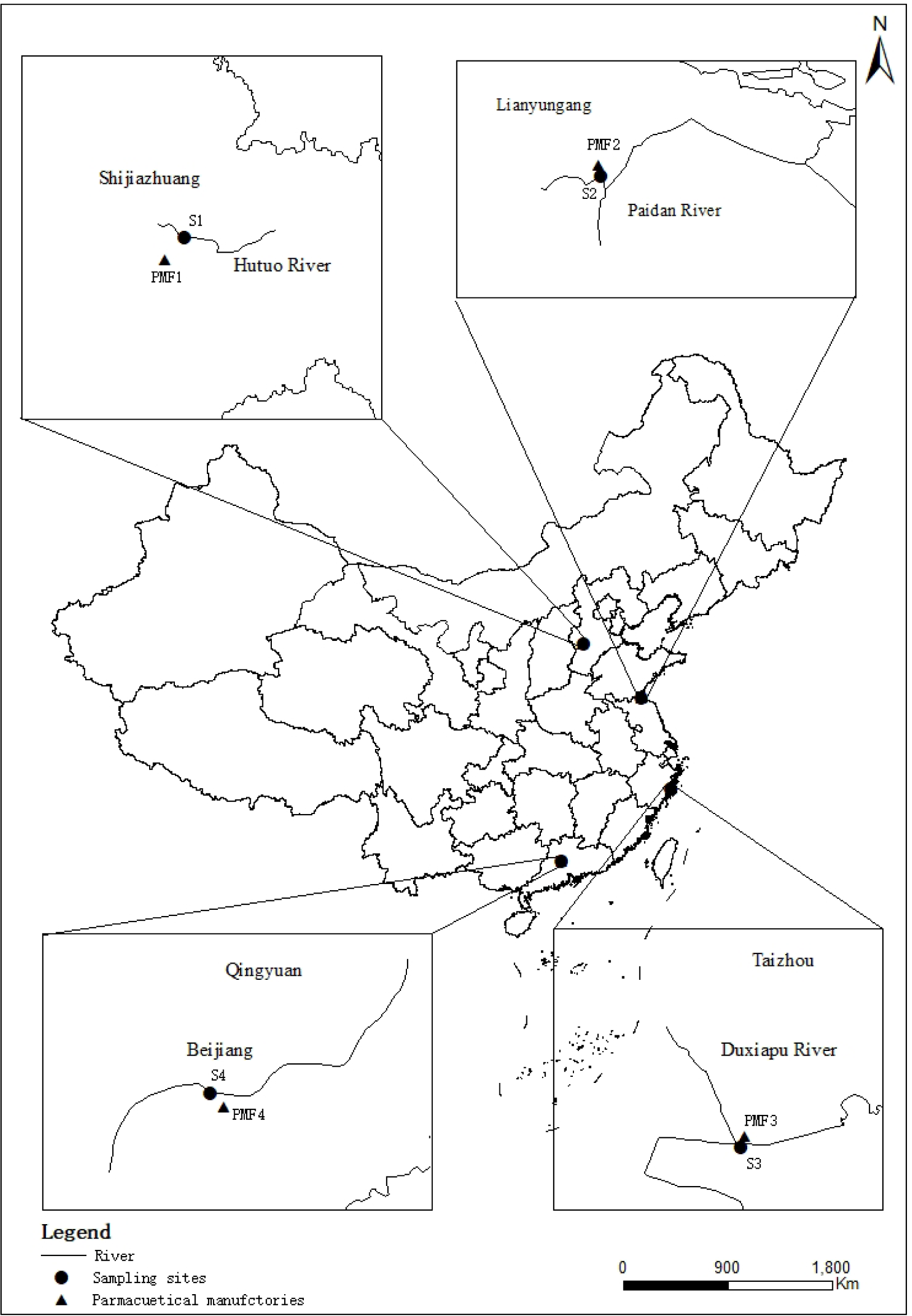
Sampling locations of the four PMFs (PMF1-PMF4) and other sites in China.

**Figure 2.**
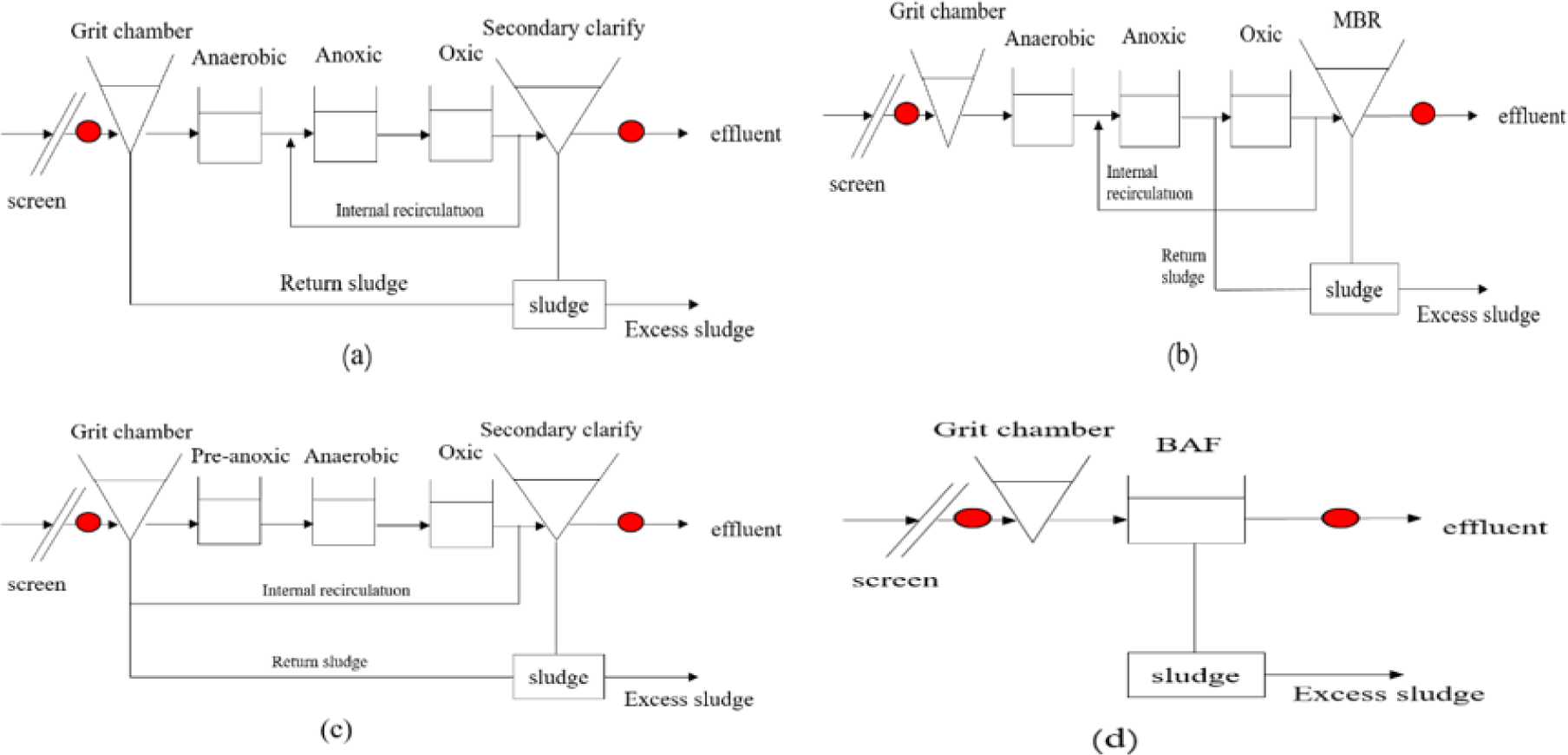
Flow charts of technological processes and sampling sites for four PMFs: (a) PMF1, A2/O, (b) PMF2, MBR, (C) PMF3, Modified A/O, and (d) PMF4, BAF.

### 2.3 Quantification of antibiotics

The concentrations of target antibiotics in wastewater samples were measured using solid-phase extraction (SPE) combined with liquid chromatograph coupled to tandem mass spectrometry (LC-MS/MS, Agilent Liquid Chromatography 1260 coupled to an AB SCIES API-4000 triple quadrupole MS) under multiple reaction monitoring (MRM) conditions with positive electrospray ionization (ESI) mode, as described in our previous studies (Huang et al., 2021) and the EPA method (Ferrer et al., 2010).

Briefly, 250 mL of wastewater samples (pH 3.0) was first spiked with isotopically labeled internal standards, including SMX-D_4_, ERY-^13^C-D_3_, TBD-D_4_, CFX-D_8_, SMZ-^13^C_6_, TMP-D_3_, LIN-D_3_. Then, the spiked samples were passed through a pre-conditioned SPE cartridge for extraction (Text S1). In the extracts, the target antibiotics were separated using a C18 column (Agilent ZORBAX Eclipse Plus, 100 mm × 2.1 mm, 1.8 μm) during LC-MS/MS analysis. The mobile-phase solutions were 0.2% formic acid and 2 mM ammonium acetate (A) and acetonitrile (B). The antibiotics were quantified in positive electrospray ionization mode. The LC gradient program for antibiotic separation and full MS/MS measurement conditions for the individual compound are reported in Table S3 in the Supplementary Materials.

### 2.4 Quality assurance and quality control (QA/QC)

Concentrations of the target antibiotics in the samples were performed using internal standard method. And the data generated from the analysis were subject to strict quality control procedures. Procedural blanks and parallel samples (one per ten samples) were inserted during all testing as a regular part of the analysis. Results in the field and procedural blanks for all analytes were below the limit of detection (LOD). The internal standard method was used for quantification with standard curves of 12 points ranging from 0.1 ng/mL to 500 ng/mL. The coefficients (R2) for all target analytes were over 0.99. All of the target antibioticswere below the LOD in all blank samples (i.e., Milli-Q water).The analytical quality parameters LOD, LOQ, and recovery values (%) are listed in Table S4. The LOD ranged from 0.16 ng/L to 1.78 ng/L, whereas LOQ ranged from 0.52 to 5.88 ng/L. Concerning the extraction methodology, the recoveries achieved for all target compounds ranged between 60% and 130 %.

### 2.5 Statistical analysis

The statistical summary of experimental data was completed in Excel 2013. The graphics were drawn using OriginPro 2021, and the correlation analysis was completed in IBM SPSS 26.

### 2.6 Risk assessment

In this study, the potential ecological risks of target antibiotics to aquatic ecosystems were assessed by calculating the risk quotients (RQs) of antibiotics in final wastewater and receiving waters (Patrolecco et al., 2015)

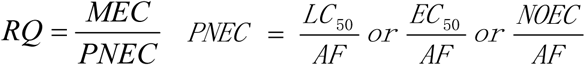

where MEC is the measured maximum environmental concentration (ng/L) of the target compound, and PNEC is the predicted no-effect concentration (ng/L) of the target compound in the water body. The information on PNECs was obtained from previously published works (Table S5). The risk was divided into four grades according to the RQ value of individual antibiotics: insignificant (RQs < 0.01), low risk (0.01 ≤ RQs ≤ 0.1), intermediate risk (0.1 ≤ RQs ≤ 1.0), and high risk (RQs > 1.0) (Hu et al., 2018; Martínez-Alcalá et al., 2021).

## 3. Result and Discussion

### 3.1 Presence of antibiotics in raw influents and treated effluents from PMFs

Seventeen antibiotics were investigated in the influents and effluents of four PMFs in Hebei, Jiangsu, Zhejiang, and Guangdong provinces. Of the 17 selected antibiotics, 13 were detected in wastewater samples from all PMFs, including 5 sulfonamides (SMZ, SD, SMX, SPD, TMP), 4 macrolides (CTM, ERY, LIN, ROX), 3 fluoroquinolones (CIP, NOR, OFL) and 1 tetracycline (MTC). Their detection frequencies in all samples are shown in Table S6. ERY and OFL were the antibiotics of the highest detection frequency (75%). TMP, LIN, and ROX occurred with a detection frequency of more than 50%. Among the wastewater samples investigated, the influents had a higher detection frequency (76.5%) than effluents (64.7%). Overall, the high detection frequencies suggested the widespread existence of antibiotics in the PMFs from Hebei, Jiangsu, Zhejiang, and Guangdong, China.

The average concentrations of SAs, FQs, MLs, and TCs in the wastewater samples are summarized in Table 1. The total concentrations of all antibiotics (the sum of all the antibiotics in a certain wastewater sample) ranged from 57.03 to 726.79 ng/L in all wastewater samples.

**Table 1.** The concentration range, median concentration, and the detectionfrequencies of antibiotics in wastewater samples (ng/L)(n=16)

| Analyte |  | Influent |  | Effluent |  |
| --- | --- | --- | --- | --- | --- |
|  |  | Range(median) | Detection<br>frequency(%) | Range(median) | Detection<br>frequency(%) |
| SAs | Sulfadiazine | nd-2.92(nd) | 25 | nd-3.71(nd) | 25 |
|  | Sulfamethazine | nd-2.5(nd) | 18.8 | nd | 0 |
|  | Sulfamethoxazole | nd-159.15(nd) | 25 | nd-24.02(nd) | 25 |
|  | Sulfapyridine | nd-39.21(nd) | 25 | nd-3.77(nd) | 25 |
|  | Trimethoprim | nd-39.91(8.43) | 75 | nd-16.81(nd) | 75 |
| MLs | Clarithromycin | nd-3.82(nd) | 25 | nd-45.26(nd) | 25 |
|  | Erythromycin-H<sub>2</sub>O | 9.01-41.31(19.24) | 100 | nd-100.53(31.81) | 56.3 |
|  | Lincomycin | nd-133.05(3.1) | 50 | nd-252.33(15.3) | 50 |
|  | Roxithromycin | nd-68.26(5.59) | 25 | nd-42.04(1.94) | 50 |
| FQs | Ciprofloxacin | nd-4.93(4.78) | 50 | nd-63.87(25.5) | 25 |
|  | Norfloxacin | nd-109.01(nd) | 31.2 | nd | 0 |
|  | Ofloxacin | nd-179.03(35.12) | 75 | nd-455.71(18.77) | 75 |
| TCs | Metacycline | nd-16.87(nd) | 18.8 | nd-31.76(nd) | 25 |

nd: not detected.

And the concentrations of detected antibiotics were in the range of 125.49–403.9 ng/L in the raw influent and 57.03–726.79 ng/L in the treated effluent.

In the present study, FQs and MLs were the most abundant antibiotic classes detected in most wastewater samples, accounting for 42.8% and 38.7% of total antibiotic concentrations, respectively, followed by SAs (16.4%) and TCs (2.4%). And the total concentrations of the different antibiotic classes in all samples followed the rank order ΣFQs [not detected (nd)– 455.7 ng/L, (106.98 ± 170.32 ng/L)] > ΣMLs (2.5–252.34 ng/L, 97.39±95.08 ng/L) > ΣSAs (1.94–183.36 ng/L, 41.18 ± 63.02 ng/L) > ΣTCs (nd–31.76 ng/L, 6.08 ± 11.94 ng/L) (Table 1). The data presented in the study were several orders of magnitude lower than those reportedfrom other countries or regions (Table 2). SIM investigated the levels of antibiotics in wastewater treatment facilities of four pharmaceutical factories in Busan, South Korea; both FQs and MLs were more than three orders of magnitude higher than those in this study (Sim et al., 2011). In the wastewater treatment facilities of PMFs in Beijing, the detected mass concentrations of TCs and SAs in influents and effluents were more than six orders of magnitude higher than those in this study (J. Wang et al., 2015)..

**Table 2.** Comparison of the antibiotic concentration levels in wastewater samples between this study and previous studies

| Compound | PMF |  |  |
| --- | --- | --- | --- |
|  | This study (ng/L) | Other studies (ng/L) | Country/Regions |
| CIP | Influent: <LOQ-1.38<br>Effluent: <LOQ-33.58 | Effluent: 28,000–31,000 × 10<sup>3</sup> | Sweden (Larsson et al., 2007) |
| CTM | Influent: <LOQ-4.93<br>Effluent: <LOQ-45.26 | Effluent: 40–800 | Vietnam (Thai et al., 2018) |
| ERY-H2O | Influent: <LOQ-1.38<br>Effluent: <LOQ-33.58 | Influent: 30.0–750.0<br>Effluent: <LOQ | Tianjin, China (Hou et al., 2019) |
| LIN | Influent: nd–133.05<br>Effluent: nd–252.33 | Effluent: Up to 35,538: | Shanghai, China (Pan et al., 2020) |
| NOR | Influent: nd–109.01<br>Effluent: nd | Effluent: 390–420×10<sup>3</sup> | Sweden (Larsson et al., 2007) |
| OFL | Influent: nd–179.03<br>Effluent: nd–455.71 | Effluent: 150–160×10<sup>3</sup> | Sweden (Larsson et al., 2007) |
| ROX | Influent: nd–68.26<br>Effluent: nd–42.04 | Influent: <10–375.3<br>Effluent: <LOQ | Tianjin, China (Hou et al., 2019) |
| MTC | Influent: nd–16.87<br>Effluent: nd–31.76 | - | - |
| SD | Influent: nd–2.92<br>Effluent: nd–3.71 | Effluent: 3.0–20.0 | Croatia (Bielen et al., 2017) |
| SMZ | Influent: nd–2.5<br>Effluent: nd | Effluent: 6.7–231.0 | Croatia (Bielen et al., 2017) |
| SMX | Influent: nd–159.15<br>Effluent: nd–24.02 | Effluent: Up to 50 | Vietnam (Thai et al. 2018) |
| SPD | Influent: nd–39.91<br>Effluent: nd | - | - |
| TMP | Influent: nd–39.21<br>Effluent: nd | Effluent: 1–10 × 10<sup>3</sup> | India (Fick et al. 2009) |

Concentration\*: the range of minimum to maximum concentrations of antibiotics detected in the wastewaters.
<LOQ: below the limit of quantification (LOQ) of analytical method.
nd: not detected.
–: data were not found in literature.

#### (1) Fluoroquinolones

The FQs is the most abundant antibiotic class in all PMFs wastewater samples, with the concentrations up to 455.7ng/L. And the ofloxacin concentration was the highest among the three detected FQs and ranged from nd to 179.03 ng/L (56.89 ± 82.7 ng/L) in influents and from nd to 455.71 ng/L (121.24 ± 223.11ng/L) in effluents. Norfloxacin was the second highest, and the concentration ranged from nd to 109.01 ng/L (influent = 27.25 ± 54.51 ng/L; nd in effluent) in wastewater samples. The concentrations of ofloxacin and norfloxacin in the raw influent of this study were lower than those in PMF wastewater in Vietnam (Thai et al., 2018) and Tianjin, China (Hou et al., 2019). The concentrations of other detected antibiotics within the FQ class were much lower than those reported for Sweden (Larsson et al., 2007), but similar to those of countries that mainly import antibiotics and active ingredients.

#### (2) Macrolides

MLs were the second most abundant antibiotic groups detected in pharmaceutical plants. Erythromycin-H_2_O, leucomycin, and roxithromycin were the most frequently detected MLs in all wastewater samples; of these, the concentration of lincomycin was the highest and in the range of nd–252.3 ng/L (66.91 ± 123.83 ng/L) in influents and changed from nd–133.05 ng/L (34.04 ± 66.02 ng/L) in the effluent samples. These levels were much lower than the previously reported concentrations of lincomycin in PMF effluent samples from Shanghai (35538ng/L). Clarithromycin was occasionally detected in concentrations ranging from nd to 45.26 ng/L at the studied PMFs, which were much less contaminated than those of the wastewater influents and effluents at the PMFs in Korea and the USA (Cardoso et al., 2014). In general, the MLs were occasionally detected in the wastewater samples of PMFs. Such differences might be related to the national regulations on antibiotic usage; for example, most ML antibiotics have been banned in aquaculture in China since 2002.

#### (3) Sulfonamides

In four PMFs, five out of nine investigated SAs were detected at least once in influent and effluent samples. The order for the average concentrations of SAs in the top three was sulfamethoxazole > trimethoprim > sulfapyridine. Sulfamethoxazole was found with the highest concentration (159.15 ng/L) in influent wastewater samples. The annual consumption of sulfamethoxazole in the world was approximately 600–800 t/year, ranking at the top among prescribed antibiotics drugs.(Johnson et al., 2015; Zhang et al., 2015). As a result, a relatively high concentration of sulfamethoxazole was found in PMF effluents compared with the other SAs.

#### (4) Tetracyclines

Only metacycline was found in all wastewater samples among TCs investigated. The concentration of metacycline ranged from nd to 16.87 ng/L in influents and from nd to 31.76 ng/L in effluents. In general, TCs were occasionally detected in wastewater samples from the studied PMFs, which were much less contaminated than those of the wastewater influents and effluents at the PMFs in Korea (Cardoso et al., 2014). This difference might be due to the national regulations on antibiotic usage. Also, several TCs were not recommended for human usage due to their acute toxicity.

### 3.2 Removal of antibiotics in PMFs

Comparing the removal efficiency of each antibiotic separately, we found that the removal efficiencies varied largely from –1084.8% to 100% for the detected antibiotics in four PMFs (Table 3). Among the 17 antibiotics investigated, the highest removal efficiency was for norfloxacin, reaching 100%, which was similar to the value (93.9%) at the PMFs from Qingdao, China (Wang et al., 2021). Sulfapyridine and sulfamethoxazole were the second highest, and their average removal efficiency was 90.3% and 89.6%, respectively. The removal efficiency of sulfamethoxazole (–31.2 to 1.8%) was detected in PMFs from China (Wang et al., 2021), which was lower than that detected in the studied PMFs. The mean removal efficiency for trimethoprim was only 22.17%, which was the lowest value among the 17 antibiotics investigated. The negative removal efficiency of several antibiotics, including ciprofloxacin, clarithromycin, erythromycin-H_2_O, lincomycin, ofloxacin, and so forth, were observed in the studied PMFs, which were also reported in previous works (Cardoso et al., 2014; Larsson et al., 2007). For example, the removal efficiency of lincomycin in two PMFs in Qingdao was reported to be – 100% to 98.4% (Wang et al., 2021). The negative removal efficiency of antibiotics was observed in PMFs, which might be due to several reasons: (i) transformation of the conjugated forms into parent compounds by microorganisms (Jelic et al., 2011), (ii) release of the antibiotics enclosed in fecal particles (Göbel et al., 2007), and (iii) sampling strategies and uncertainties of sample treatments and measurements (Jelic et al., 2011; Tran et al., 2016).

**Table 3.** Overall removal efficiencies of the detected antibiotics in the wastewaters

| Analyte |  | Removal efficiencies in PMFs |  |  |  |  |
| --- | --- | --- | --- | --- | --- | --- |
|  |  | Range (%) | PMF1 | PMF2 | PMF3 | PMF4 |
| SAs | Sulfadiazine | -41.2 | nd | -41.2 | nd | nd |
|  | Sulfamethazine | nd-100 | nd | nd | 100 | nd |
|  | Sulfamethoxazole | nd-84.9 | 84.9 | nd | nd | nd |
|  | Sulfapyridine | nd-90.3 | nd | nd | 90.3 | nd |
|  | Trimethoprim | -99-100 | 64.5 | 100 | 98.9 | -99.4 |
| MLs | Clarithromycin | -1084.8 | nd | -1084.8 | nd | nd |
|  | Erythromycin-H<sub>2</sub>O | -203.2-100 | -203.2 | -143.4 | 95.1 | 100 |
|  | Lincomycin | -89.7-100 | -79.2 | nd | 100 | -89.7 |
|  | Roxithromycin | 38.4-65.3 | nd | 38.4 | nd | 65.3 |
| FQs | Ciprofloxacin | -612.9 | nd | nd | -612.9 | nd |
|  | Norfloxacin | 100 | nd | 100 | nd | nd |
| TCs | Ofloxacin | -3298.3-94.2 | nd | 94.2 | 46.6 | -3298.3 |
|  | Metacycline | -88.3 | -88.3 | nd | nd | nd |

$Removal\ efficiencies\ (\%) = \frac{C_{influent} - C_{effluent}}{C_{influent}} \times 100\%$
nd: no data

The removal efficiency for most target antibiotics varied widely and differed considerably at different PMFs in this study. For example, the removal efficiency of erythromycin-H_2_O ranged from – 203% to 100% while that of lincomycin ranged from –89.6% to 100% at four PMFs. PMF3 possessed the best removal capability for MLs (95.4%) and SAs (95.2%), and PMF2 had the best removal capability for FQs (96.2%). In this study, the removal efficiencies of antibiotics at PMF1, 2, and 3 (average removal efficiency = 50.62%) with the second treatment process of cyclic activated sludge technology(CAST)were obviously higher than those at PMF4 (–304.4%). The total concentration of target antibiotics in effluents was higher than that in influents at PMF4, which could be attributed to different treatment techniques (AS and CAST), operational parameters (hydraulic retention timeand temperature), and so forth.

### 3.3 Occurrence of the selected antibiotics in receiving waters

The antibiotics were widely detected in the water samples from the receiving waters at the level of nanogram per liter. As shown in Table 3, among 17 selected antibiotics, 13 were detected in 4 surface water samples, including 6 SAs (sulfadiazine, sulfadimethoxine, trimethoprim, sulfamethazine, sulfamethoxazole, and sulfamonomethoxine), 4 MLs (clarithromycin, erythromycin-H_2_O, lincomycin, and roxithromycin), 2 FQs (ciprofloxacin and ofloxacin), and 1 TC (methacycline). The total concentration range of 13 antibiotics in surface water samples was 27.88–353.98 ng/L. As shown in Figure 4, it was obvious that MLs had the highest concentration, with an average concentration of 152.46 ng/L, followed by SAs, QNs, and TCs, with an average concentration of 23.17, 2.41, and 1.15 ng/L, respectively. The average concentration of these four MLs was 86.55 ng/L, 32.09 ng/L, 22.84 ng/L, and 10.98 ng/L, respectively. In surface water samples, erythromycin-H_2_O (100%) was present with the highest detection frequency among four MLs; the detection frequency of clarithromycin, lincomycin, and roxithromycin was more than 50% (Table 4). Studies have shown that erythromycin-H_2_O and roxithromycin are the most commonly detected MLs in the environment of our country (Yang et al., 2016), while clarithromycin is the most common MLs in Europe and Canada (McArdell et al., 2003), indicating that different types of antibiotics were used in each country and region. In addition, the pH of surface water samples was usually between 7.5 and 8.5. MLs were stable under neutral conditions where they were not easily decomposed (Lai and Lin, 2009), which might be another reason for the high detection concentration of such antibiotics.

**Table 4.** Concentrations of antibiotics detected in the receiving waters of PMFs

279 *nd: not detected.*
| Compounds | Frequency (%) | Concentration (ng · L<sup>-1</sup>) |  |  |  |
| --- | --- | --- | --- | --- | --- |
|  |  | Med. | Max. | Min. | Ave. |
| CTM | 0.5714 | 32.25 | 119.56 | nd | 32.09 |
| ERY | 1.0000 | 73.92 | 237.49 | 11.4400 | 86.55 |
| LIN | 0.7143 | 8.56 | 105.06 | nd | 22.84 |
| OFL | 0.7143 | 2.62 | 5.98 | nd | 3.18 |
| ROX | 0.7857 | 24.53 | 30.3900 | nd | 10.98 |
| SCP | 0.4286 | 7.145 | 17.7600 | nd | 8.43 |
| MTC | 0.2500 | 5.9100 | 6.4000 | nd | 5.87 |
| SDZ | 0.4286 | 1.11 | 1.54 | nd | 1.43 |
| SDM | 0.2143 | 2.56 | 2.78 | nd | 2.57 |
| SMZ | 0.7143 | 1.43 | 1.83 | nd | 1.69 |
| SMX | 1.0000 | 4.4700 | 59.99 | nd | 13.91 |
| SMM | 0.4286 | 4.48 | 9.16 | nd | 4.745 |
| TMP | 0.7857 | 2.1600 | 5.2000 | nd | 3.10 |

Moreover, sulfamethoxazole was found with the highest detection frequency among SAs, and its average concentration was 13.91 ng/L. The concentration of sulfamethoxazole in the water environment was higher than that of other SAs, which was closely related to its large-scale use in treating bacterial infections (Göbel et al., 2005). Ofloxacin was found with the highest detection frequency among two FQs, and its average concentration was 2.03 ng/L. Compared with macrolides and sulfonamides antibiotics, the low detection concentration of FQs might be related to their easy adsorption on the surface of solid particles and easy photolysis in surface water. The detection frequency of TCs in the surface water samples in this study was only 25%, and the average concentration was 1.15 ng/L. Indeed, the TCs and MLs administrated by humans were nearly two times higher than the SA antibiotics in China (Zhang et al., 2015). The low detection frequencies and concentrations of TCs might be attributed to different medical prescription patterns in different cities and regions.

In addition, different levels of pollution were detected in four sampling points of surface waters around the PMFs (Fig. 3). The highest contamination level of all the selected antibiotics was found in the receiving water of PMF3 (S3), followed by the receiving water of PMF2 (S2). However, the lowest level of pollution was found in the Hutuo river (S1), which was the receiving water of PMF1.

**Figure 3.**
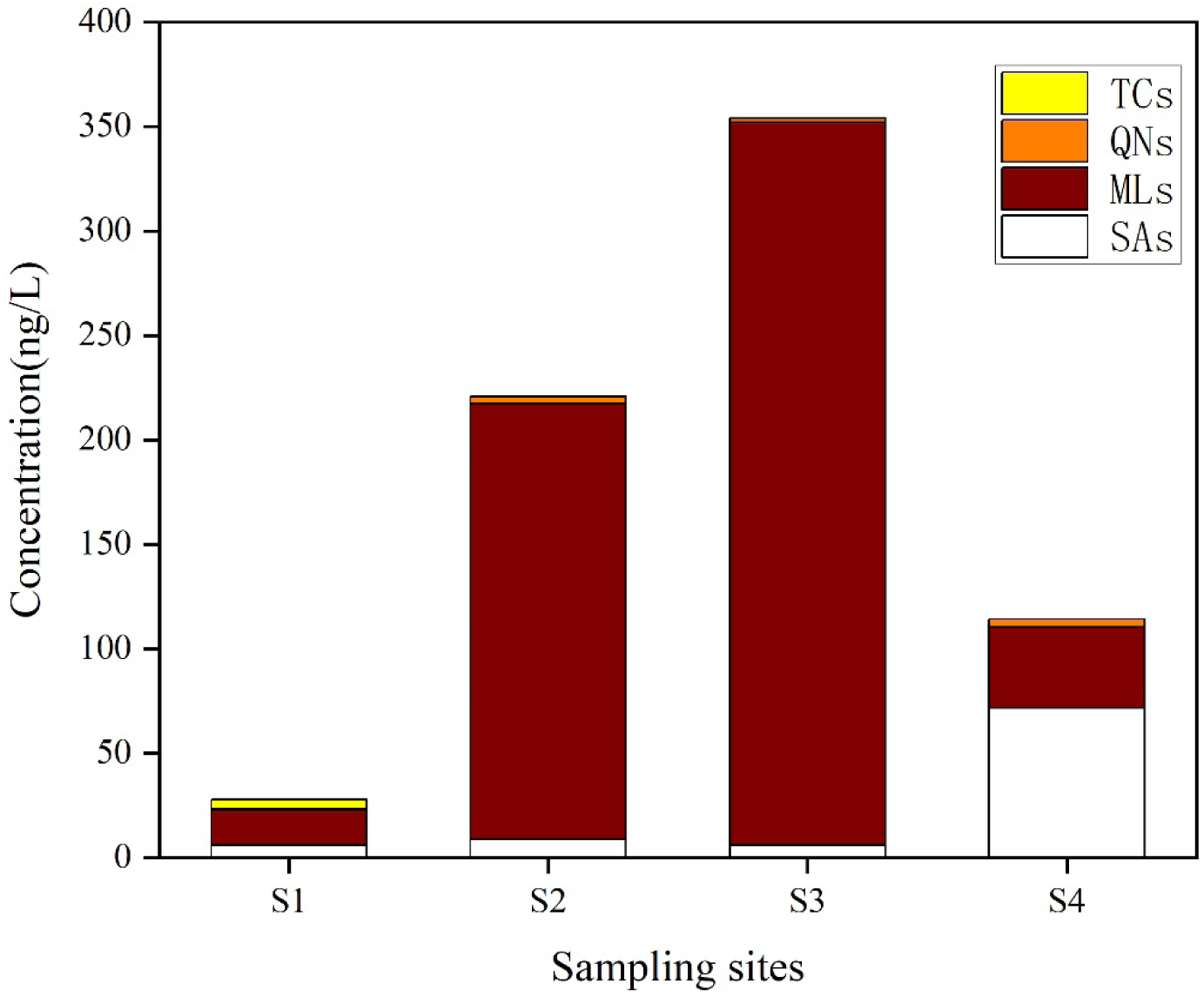
Concentrations of detectable antibiotics from four categories in the sampling sites from the receiving waters of four PMFs.

Antibiotics with a detection frequency of >70% and an average concentration of >5.00 ng/L in this study were selected to compare surface waters at home and abroad. The target antibiotic concentrations in the waters around PMFs were comparable to those in other water environments.

### 3.4 Interrelationships between selected antibiotics in effluents and receiving river

PMFs could not remove all antibiotics from influents, thus releasing antibiotics into the environment and then spreading in rivers. In general, the concentrations of antibiotics in receiving waters were lower than those in PMF effluents. A Pearson correlation analysis between the absolute concentrations of antibiotics in PMF effluents and their nearby downstream sampling sites was carried out to understand the interrelationship between antibiotics in PMFs and their receiving waters. Significant correlations were observed between PMF2 and S2 (*r* = 0.783, *P* = 0.016), and PMF1 and S1 (*r* = 0.792, *P* = 0.018), indicating that the emissions of these two PMFs might directly affect the content of antibiotics in receiving water bodies. No correlation was observed between PMF4 and S4, and PMF3 and S3. In PMF4 and its receiving waters, ciprofloxacin and sulfonamide antibiotics were detected only in the receiving river; sulfonamides were the main compounds, reflecting the many applications of sulfonamides in this field. Intensive aquaculture and poultry fishing activities might be a major source of sulfonamides. In PMF3 and its surrounding waters, antibiotics such as clarithromycin and sulfamethoxazole were detected only in the samples of receiving river, indicating that these antibiotics might not come from the wastewater of PMFs. The antibiotics in the receiving rivers might also come from other pollution sources, such as urban STPs, rural wastewater, and so forth.

### 3.5 Ecological risk assessment

Discharge from PMFs has been identified as the main point source of antibiotics in the aquatic environment, which may pose potential ecological risks to aquatic organisms as well as potential risks to the food chain (Wang et al., 2021). Antibiotics discharged into the environment have become a special environmental selection pressure, making the microorganisms carrying antibiotic resistance genes resistant and more likely to survive (Yan et al., 2014). At the same time, antibiotic resistance genes replicate and spread in the environment along with the reproduction of microorganisms, posing a serious threat to human health and ecological security. Thus, the environmental risk assessment of antibiotics in the aquatic environment is necessary. The calculated RQs of antibiotics for three aquatic organisms (algae, invertebrates, and fish) are summarized in Table S5. Algae are the most sensitive organisms in the aquatic environment to these antibiotics, which can be confirmed by other researches. By calculating RQ values in the effluent of PMFs and their receiving waters, it is concluded that the wastewaters after emission from the PMFs definitely present ecological risks. Erythromycin-H_2_O, lincomycin, ofloxacin, and sulfamethoxazole are all potential threats, indicating that they exert a relatively highly acute or chronic toxicological risk to aquatic organisms. As shown in Figure 4, ofloxacin and lincomycin pose a high risk to algae. They showed a high ecological risk for algae in effluents of PMF4 with RQ values of 21.7 and 3.6, respectively. The discharge of wastewater from the effluent of PMFs into their receiving waters certainly causes the contamination of the aquatic environment.

**Figure 4.**
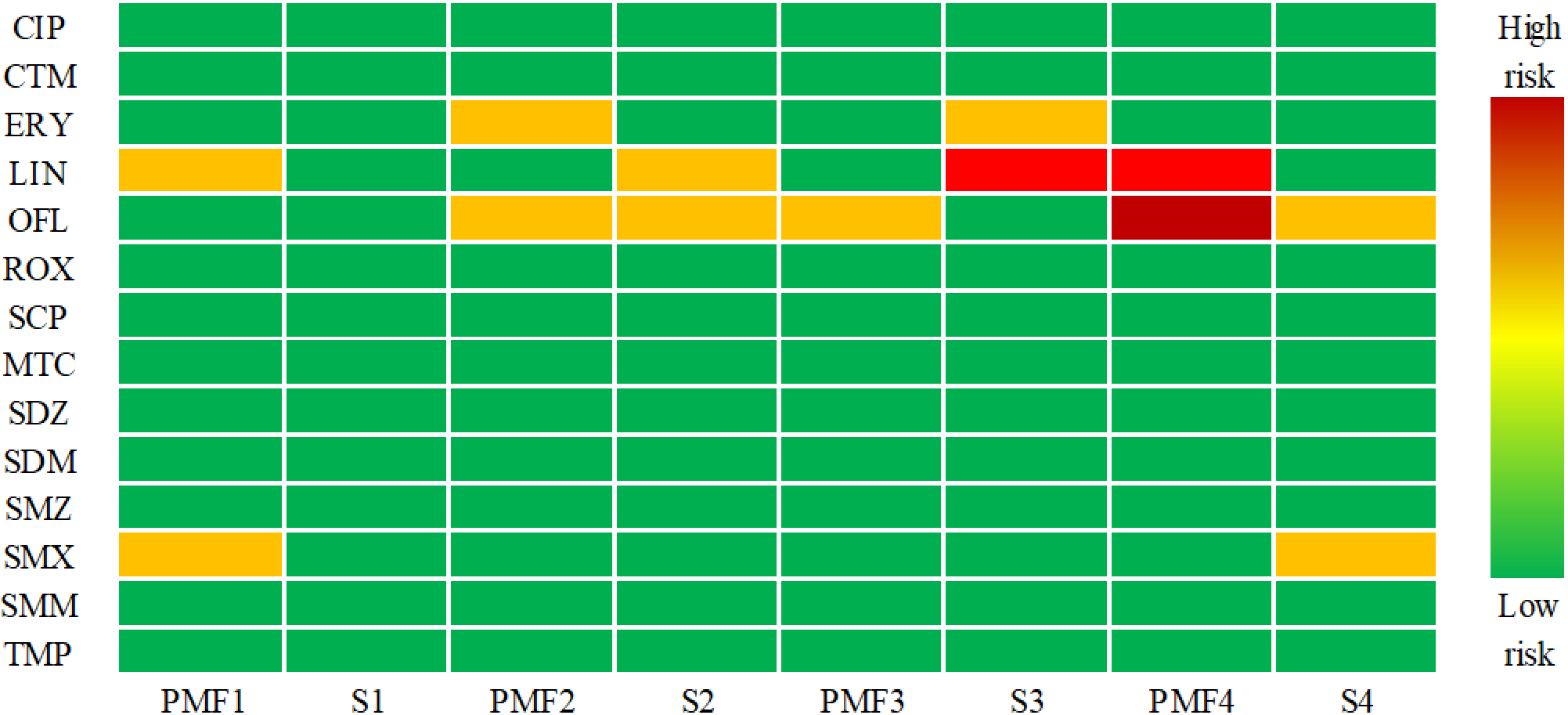
RQs for antibiotics in water samples in the PMFs and receiving waters.

The environmental risk of wastewater significantly decreases due to the dilution and sorption effect when the effluent is discharged into receiving waters. However, the risk posed by lincomycin and erythromycin-H_2_O in the receiving water (S3) is higher than that in the effluent of PMF3, which can be attributed to other pollution sources, such as sewage wastewater treatment plants and livelock farms. Among the detected sulfonamides, only sulfamethoxazole posed a medium ecological risk to algae in the effluent of PMF1 and the receiving water of PMF4. However, other sulfonamides such as sulfachlorpyridazine, sulfadiazine, sulfadimethoxine, sulfamethazine, sulfamonomethoxine, and trimethoprim posed no ecological risk to algae in all sampling sites. Among the other detected antibiotics, ciprofloxacin, clarithromycin, roxithromycin, and methacycline also posed no ecological risk in all sampling sites. Overall, 100% of the effluent samples from four typical PMFs posed medium to high ecological risk due to the high concentration of erythromycin-H_2_O, lincomycin, and ofloxacin. Although the antibiotics undergo a certain degree of dilution when they are discharged into surface water, antibiotic contamination occurs in the receiving waters due to the continuous discharge of wastewater. The environmental risk of most antibiotics is low. However, the long-term drainage into aquatic ecosystems may lead to negative effects and thus should be tested. Moreover, the mixed effect caused by the interaction of different antibiotics may be more significant than the individual effect (Leung et al., 2012). Therefore, further investigation is required.

## 4 Conclusions

Our findings indicated that the target antibiotics were frequently detected in the typical pharmaceutical plants (inffluents and effluents) and their receiving water bodies in China. The discharge of the pharmaceutical plants may directly influence the receiving water through the pearson correlation analysis. Finally, the environmental risk analysis showed that lincomycin and ofloxacin could pose a risk both in effluents and their receiving waters. In the future, more attention should be place on the seasonal variation and continuous long-term monitoring of antibiotics.

## Data Availability

All relevant data are within the manuscript and its Supporting Information files.

## Acknowledgments

This study was financially supported by the National Key Research and Development Program of China (2018YFC1801505), the National Science Foundation for Distinguished Young Scholars of China [42107468], the Guangzhou Municipal Science and Technology Project [201804010193, 202240070007], and the Special Basic Research Fund for Central Public Research Institutes of China [PM-zx097-202104-089, PM-zx703-202204-179]. We also thank the anonymous referees for their thorough review and constructive comments.

